# EEG Biomarkers for Rett Syndrome: A Comparative Analysis of Machine Learning and Deep Learning with Explainable AI

**DOI:** 10.64898/2026.10.21.26366077

**Authors:** Megha Gupta, Changhong Jin, Alejandro Lopez Valdes, Chiara Bosetti, Gemma Marinella, Bartolini, Roberta Battini, Ruihai Dong, Daniela Tropea

## Abstract

Rett syndrome (RTT) is a rare neurodevelopmental disorder characterised by abnormal cortical activity, which can be captured using electroencephalography (EEG). Although machine learning (ML) has shown promise for diagnosing several neurological conditions, comprehensive evaluation of classical ML and deep learning (DL) methods, together with physiologically meaningful explanations, remains limited for RTT. This study analyses resting-state, eyes-closed EEG recordings from RTT patients and healthy controls using both power spectral density (PSD)-based ML and end-to-end DL frameworks. Classical ML models, including Support Vector Machines, Random Forest, and XGBoost, were trained on PSD features, while EEG-specific DL architectures, including ShallowConvNet, DeepConvNet, EEGNet, and EEG Conformer, were trained directly on preprocessed EEG signals. Subjectwise cross-validation was used to avoid data leakage, with balanced accuracy as the primary performance metric because it equally accounts for sensitivity and specificity. Random Forest achieved the best performance among the classical ML models, whereas EEGNet achieved the highest DL performance. An ablation study indicated that 4-second EEG epochs offered the optimal trade-off between model performance and robustness. Model interpretability was investigated using SHAP for Random Forest and occlusion sensitivity analysis for EEGNet. Both methods consistently identified delta and theta oscillations, along with centro-frontal brain regions, as key contributors to RTT classification. These findings show that both PSD-based machine learning and end-to-end deep learning methods can reliably identify RTT and highlight common neurophysiological biomarkers, offering an interpretable framework for EEG-based diagnosis and biomarker discovery for Rett Syndrome.

## 1. Introduction

Rett syndrome (RTT) is a severe neurodevelopmental disorder, affecting approximately 1 in 10,000 live female births [1, 2]. It was first described by Andreas Rett in the 1960s and is primarily caused by mutations in the Methyl-CpG-binding protein 2 (MECP2) gene [3]. RTT is characterised by an initial period of normal development, followed by a progressive deterioration of motor and cognitive skills, seizures, and respiratory problems, with symptoms generally appearing between 6 and 18 months of age. Diagnosing these rare diseases depends on clinical observations and genetic tests [4]. While ongoing research aims to better understand their underlying mechanisms [5, 6] and develop potential treatments, the variability in their presentation and rarity complicate patient classification, management, and testing of new therapies [7, 8, 9, 10, 11].

Electroencephalography (EEG) has emerged as a valuable tool for investigating the neurophysiological abnormalities associated with RTT [12, 13, 14, 15]. Previous studies have reported alterations in background activity, including reduced posterior alpha rhythms [16], and increased slow-wave oscillations [14, 17]. EEG has also been used to identify phenotypic or genotypic differences [12], to assess the potential efficacy of experimental treatments such as the impact of Insulin-Like growth factor 1 (IGF1) on network formation in responders versus non-responders [13], and to measure disease severity [18]. These findings suggest that EEG may provide objective biomarkers to support diagnosis, prognosis, patient stratification, or monitor disease progression.

Traditional EEG research primarily identifies disease-related abnormalities by comparing specific EEG features between patients and healthy controls (HC) using statistical tests such as Kruskal-Wallis tests, Mann-Whitney U tests, or Spearman correlation [12, 14, 15]. These methods analyse features separately and might miss the complex relationships among multiple variables in high-dimensional EEG data. Machine learning (ML) overcomes this by modelling multiple features and their interactions simultaneously, enabling the development of data-driven classifiers for automated disease detection.

Recent research shows that combining EEG biomarkers with ML can effectively characterise RTT. Resting-state EEG coherence measures, classified with a Support Vector Machine (SVM), predicted responses to IGF-1 treatment with perfect accuracy, underscoring their utility in patient stratification and clinical trial recruitment [13]. Spectral analysis of resting-state EEG revealed increased fronto-central theta activity, reduced alpha power, and heightened gamma oscillations in RTT patients. These neural signatures distinguish RTT from typically developing children with high accuracy (AUC *≈* 0.92) using a Random Forest model [16]. Furthermore, auditory event-related potentials (ERPs) analysed with both linear and nonlinear ML techniques achieved AUCs up to 0.94, with changes in the P2-N2 response serving as a key marker [19]. Overall, these findings highlight that EEG contains valuable diagnostic information for RTT, but most studies rely on handcrafted features with classical ML models. The potential of modern deep learning (DL) architectures tailored for EEG analysis in RTT remains underexplored, partly due to the disorder’s rarity and limited data availability. As a result, it remains unclear whether handcrafted features or end-to-end deep learning models applied directly to raw EEG data offer the most effective and interpretable approach for RTT classification.

To address these gaps, this study provides a detailed comparison of classical ML and DL methods for automatically classifying individuals with RTT and HC using resting-state, eyes-closed EEG recordings. To our knowledge, it is the first to systematically evaluate power spectral density (PSD)-based classical ML models and end-to-end DL models that operate directly on preprocessed EEG signals within a unified, subject-wise evaluation framework. In addition to assessing diagnostic performance, the study aims to identify physiologically meaningful, interpretable EEG biomarkers by applying explainable Artificial Intelligence (XAI) techniques to explore the spatial and spectral features that influence model predictions. Finally, by comparing biomarkers derived from both approaches, this work investigates whether they reveal common neurophysiological signatures of RTT, offering insights into cortical abnormalities and supporting the development of reliable, interpretable EEG biomarkers suitable for AI-assisted diagnosis.

## 2. Experimental Design and Methodology

### 2.1 Participants

This study involved two participant groups: individuals with Rett Syndrome and non-matched healthy controls. All participants were enrolled after approval from the Pediatric Ethics Committee of the Tuscany Region (CEPR). Informed written consent was obtained from the legal guardians or caregivers prior to data collection. This was part of the ongoing prospective observational study, BIRS (Biomarkers in Rett Syndrome), with CEPR registration number 99/2020, dated 28th April 2020.

Participants with RTT aged 0-18 years were recruited during clinic visits at IRCCS Stella Maris Foundation in Pisa, Italy. They were diagnosed with RTT during the developmental period, with MECP2 mutations, both classic and atypical, confirmed by MECP2 gene sequencing (NGS). The study involved annual follow-ups and the collection of standardised parameters and EEG data. Subjects with impairments that may have compromised the execution of the evaluation and monitoring procedures required by the protocol (motor and visual function assessment, EEG registration) were excluded from the study.

HC participants aged 0-18 years were recruited from the same geographic region. They had no personal history of neurological, neurodevelopmental, or psychiatric disorders and were not currently taking psychoactive, anticonvulsant, or centrally acting medications. They were excluded if they had any history of developmental delay, motor impairment, or epilepsy, or were unable to cooperate during EEG recordings.

The demographic and clinical details of all participants are summarised in Table 1.

**Table 1:** Demographic and clinical characteristics of study participants. Values are presented as mean *±* standard deviation unless otherwise specified. HC = Healthy Controls; RTT = Rett Syndrome; N/A = Not Applicable.

| Characteristic | HC (n = 9) | RTT (n = 20) |
| --- | --- | --- |
| **Demographics** |  |  |
| Age (years), mean ± SD | 17.12 ± 4.82 | 8.75 ± 3.29 |
| Age range (years) | 6.8 - 24.3 | 5.4 - 17.4 |
| Sex, female, *n* (%) | 4 (44.44%) | 20 (100%) |
| Age at diagnosis (years)<sup>a</sup>, mean ± SD | N/A | 2.63 ± 0.58 |
| **Clinical Features (RTT only)** |  |  |
| International Scoring System (ISS) severity, *n* (%)<sup>b</sup> | N/A |  |
| Mild | N/A | 7 (35%) |
| Moderate | N/A | 4 (20%) |
| Severe | N/A | 7 (35%) |
| History of seizures, *n* (%) | N/A | 7 (35%) |
| Sleep disturbance<sup>a</sup>, *n* (%) | N/A |  |
| Normal sleep patterns | N/A | 9 (45%) |
| Minor disturbances | N/A | 8 (40%) |
| Significant disturbances | N/A | 1 (5%) |

<sup>a</sup> Data for six participants was missing.
<sup>b</sup>Data was available for 18 RTT participants. For participants with multiple ISS assessments recorded at different time points, the latest severity level was used.

### 2.2 EEG Acquisition Protocol

EEG data were collected using the Brain Quick system by Micromed (v1.02.0002) [20], a widely used clinical-grade EEG platform. Before recording, 20 cup electrodes were placed on the scalp according to the International 10/20 System: eight over the right hemisphere (Fp2, F4, F8, C4, P4, T4, T6, O2), eight over the left (Fp1, F3, F7, C3, P3, T3, T5, O1), and four along the midline (Fz, Cz, Pz, Oz) [21], secured with adhesive or conductive paste. A reference electrode(vertex electrode, Cz), a ground electrode (Fpz), and two polygraphic channels measuring deltoid muscle activity (delt.Sx and delt.Dx) were also attached. The impedance of each electrode was verified to be below 10 *k*Ω before recording; if needed, additional conductive gel was applied to meet this threshold, ensuring signal quality across channels [22]. All signals were sampled at 256 Hz and recorded using a superior and inferior longitudinal bipolar montage [23], as shown in Fig. 1. The montage for the right hemisphere included Fp2-F4, F4-C4, C4-P4, P4-O2, Fp2-F8, F8-T4, T4-T6, and T6-O2; for the left, Fp1-F3, F3-C3, C3-P3, P3-O1, Fp1-F7, F7-T3, T3-T5, and T5-O1; and along the midline, Fz-Cz, Cz-Pz, and Pz-Oz. Although EEG signals were initially acquired using a longitudinal bipolar montage during the clinical session, the data were re-referenced to a common reference montage with BrainQuick software (Micromed) before sharing. The re-referenced data, with all channels referenced to a single common electrode, were then provided to the research team for analysis in the European Data Format (.edf).

**Figure 1:**
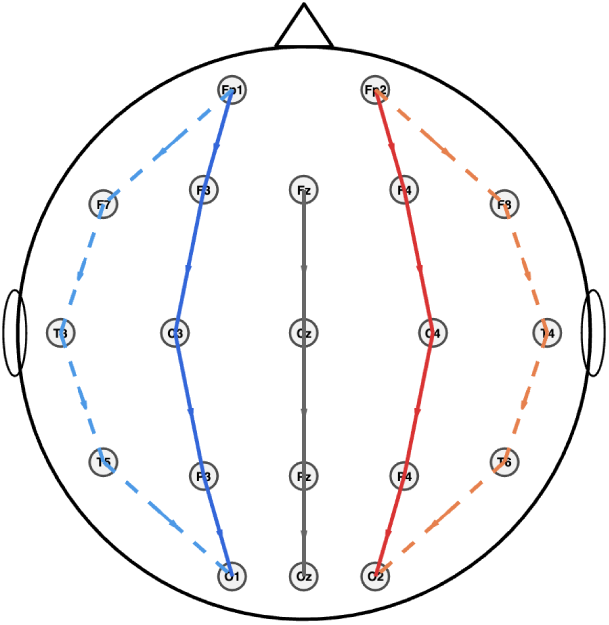
EEG channel montage used in the study. The plot shows the retained EEG channels common to both Rett syndrome and healthy control recordings. Channel locations were assigned using standard scalp electrode coordinates in EEGLAB.

Each session followed a standardised multi-phase protocol, with the awake period lasting about 20-30 minutes. Recordings began with a resting-state baseline under Eyes Closed (EC) and Eyes Open (EO) conditions, with transitions manually managed by placing a sheet of paper in front of participants’ eyes as needed. When clinically feasible, this was followed by approximately three minutes of hyperventilation to assess EEG changes induced by activation. Intermittent photic stimulation (IPS) was then applied, delivering flash trains at 1, 5, 10, 15, 18, 25, and 30 Hz to assess visual cortical reactivity and detect photoparoxysmal responses. Session annotations combined automated event marking, in which the Brain Quick system (version 1.02.0002) timestamped stimulus onset, with manual entries by trained EEG technicians for behavioural states (EC, EO, sleep stages, awakening), following a standardised protocol to maintain consistency across recordings.

### 2.3 Preprocessing Pipeline

EEG signals are often contaminated by eye blinks, body movements, environmental noise, abnormal signal fluctuations, power-line interference, and baseline drift, which can significantly reduce the reliability of EEG-based assessments [24]. To address these issues, standard preprocessing steps are applied, including noise filtering, channel removal, re-referencing, and artefact identification and removal [25].

We performed the EEG preprocessing in MATLAB R2024b [26] using custom scripts and the EEGLAB toolbox (version 2025.0.0) [27]. The same preprocessing pipeline was applied to both RTT and HC datasets to ensure methodological consistency across groups.

EEG data were imported in European Data Format (.edf) using the EEGLAB toolbox. To ensure uniformity across datasets, only the 20 EEG channels common to all RTT and HC recordings were retained, and all other EEG and non-EEG channels were removed. Signals were band-pass filtered between 1 and 45 Hz using a zero-phase finite impulse response (FIR) filter (pop_eegfiltnew) to remove slow baseline drift and high-frequency noise while preserving EEG frequency bands of interest (delta, theta, alpha, beta, and the lower part of the gamma band) [28, 29]. The recordings were then re-referenced to a common average and visually inspected to ensure signal quality. No flat or unusable channels were detected, so no further channel rejection was required. Independent Component Analysis (ICA) was not performed because the limited number of channels (20) could lead to unstable component estimates and reduced interpretability [30].

Resting-state eyes-closed segments were automatically extracted using a custom MATLAB script that utilised EEGLAB functions. Event labels "Eyes Closed" and "Eyes Opened" identified in the original .edf metadata guided the selection. EEG samples corresponding to these events were extracted while maintaining the original clinical annotation boundaries. This process resulted in 189 EC segments from RTT recordings and 67 from HC recordings. Although RTT had more EC segments, their duration distributions were similar (RTT: 9.09 ± 4.78 s; HC: 9.61 ± 4.29 s), suggesting comparable artefact-free resting-state recording lengths across groups (Figure 2).

**Figure 2:**
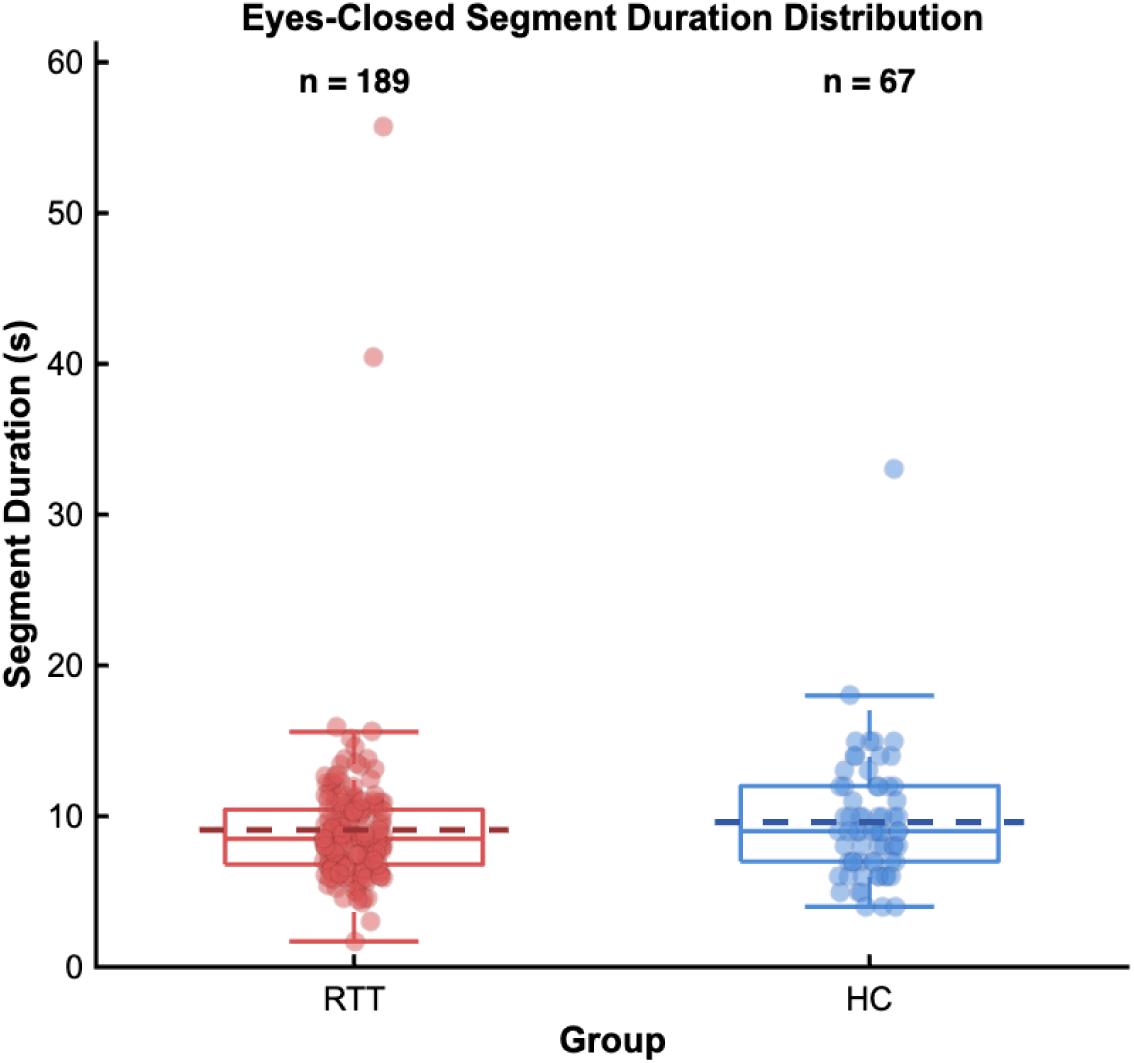
Distribution of eyes-closed segment durations. Box plots display median (central line), interquartile range (box), and range (whiskers) for Rett syndrome (RTT, red, n = 189 segments) and healthy control (HC, blue, n = 67 segments) groups. The dotted line represents the mean duration.

When multiple EC segments were present in a recording, they were kept as separate segments rather than combined to avoid artificial spectral discontinuities at the boundaries. Each EC segment was then split into nonoverlapping 4-second epochs, yielding 337 RTT and 136 HC epochs for further analysis. The analysis focused on resting-state EC EEG because it provides a consistent measure of intrinsic brain activity without task-related influences [31]. The eyes-closed state typically enhances oscillatory activity in the alpha band (8-13 Hz), which is one of the most prominent rhythms in HC [32]. Deviations from this pattern, such as decreased alpha power or increased slow rhythms (e.g., delta and theta activity), have been observed in RTT and may serve as useful spectral biomarkers [14]. Additionally, limiting the analysis to EC segments avoided potential confounds caused by stimulus-driven procedures in parts of the recordings, such as photic stimulation or hyperventilation, which can produce frequency-specific responses unrelated to resting brain activity.

### 2.4 Frequency-Domain Feature Extraction

To characterise the spectral properties of the extracted eyes-closed EEG segments, we computed power spectral density (PSD) features for each channel using Welch’s method [33, 34, 35]. The EEG signals were segmented into overlapping 512-sample windows with 50% overlap. A Hamming window was applied to reduce spectral leakage, and the PSD was estimated using a 1024-point Fast Fourier Transform (FFT) [36]. The resulting periodograms were averaged to obtain stable PSD estimates for each channel. Rather than grouping spectral power into predefined frequency bands, we retained the full PSD to preserve detailed spectral information. The 1024-point FFT yielded 513 non-redundant frequency bins. PSD features from all 20 channels were concatenated into a 10,260-dimensional vector (20 × 513) per epoch and used as input to classical machine learning models.

### 2.5 Machine Learning Algorithms

The models evaluated to train classical ML classifiers were Support Vector Machine [37], Logistic Regression (LR) [38], k-Nearest Neighbors (kNN) [39], Decision Tree (DT) [40], Random Forest (RF) [41], Gradient Boosting (GB) [42], and eXtreme Gradient Boosting (XGBoost) [43]. These models span a broad range of classification approaches commonly used in EEG-based diagnostic research [44, 45, 46].

Before training the models, feature normalisation was applied for classifiers sensitive to the scale of input variables. Specifically, z-score standardisation was performed on models that use distance or marginal optimisation, such as SVM, kNN, and LR [47]. In this process, each feature was centred by subtracting its mean and scaled by its standard deviation, ensuring all features contributed equally and improving numerical stability. In contrast, tree-based classifiers such as DT, RF, GB, and XGBoost did not require feature scaling because these algorithms partition the feature space using threshold values, making them inherently insensitive to differences in feature scales [47].

### 2.6 Deep Learning Architectures

In addition to the PSD-based machine learning analysis, deep learning models were trained directly on the preprocessed EEG time series, without relying on handcrafted features. Each input was represented as a multi-channel EEG matrix of size *C × T*, where *C* denotes the number of EEG channels and *T* the number of time samples.

To evaluate end-to-end EEG learning, four state-of-the-art architectures were examined: ShallowConvNet, DeepConvNet, EEGNet, and EEG Conformer. These models represent diverse design philosophies, ranging from physiologically inspired shallow convolutional neural networks (CNNs) to transformer-based architectures. ShallowConvNet [48] was designed specifically for EEG decoding, replicating traditional EEG signal processing by learning band-power features via temporal and spatial convolutions. DeepConvNet [48] builds on this by utilising a deeper convolutional structure to capture more complex, hierarchical temporal and spatial features. EEGNet [49] is a compact, EEG-tailored architecture that uses depthwise and separable convolutions to efficiently extract temporal and spatial information while keeping the number of trainable parameters low, making it suitable for smaller EEG datasets. EEG Conformer [50] uses convolutional-based feature extraction with transformer-based self-attention to model both local EEG patterns and long-range temporal relationships. Collectively, these architectures provide a comprehensive comparison across shallow, deep, compact, and attention-based deep learning methods for RTT classification.

### 2.7 Model Training and Evaluation

Figure 3 presents the unified evaluation framework comparing classical ML methods using PSD features with end-to-end DL models applied directly to preprocessed EEG signals.

**Figure 3:**
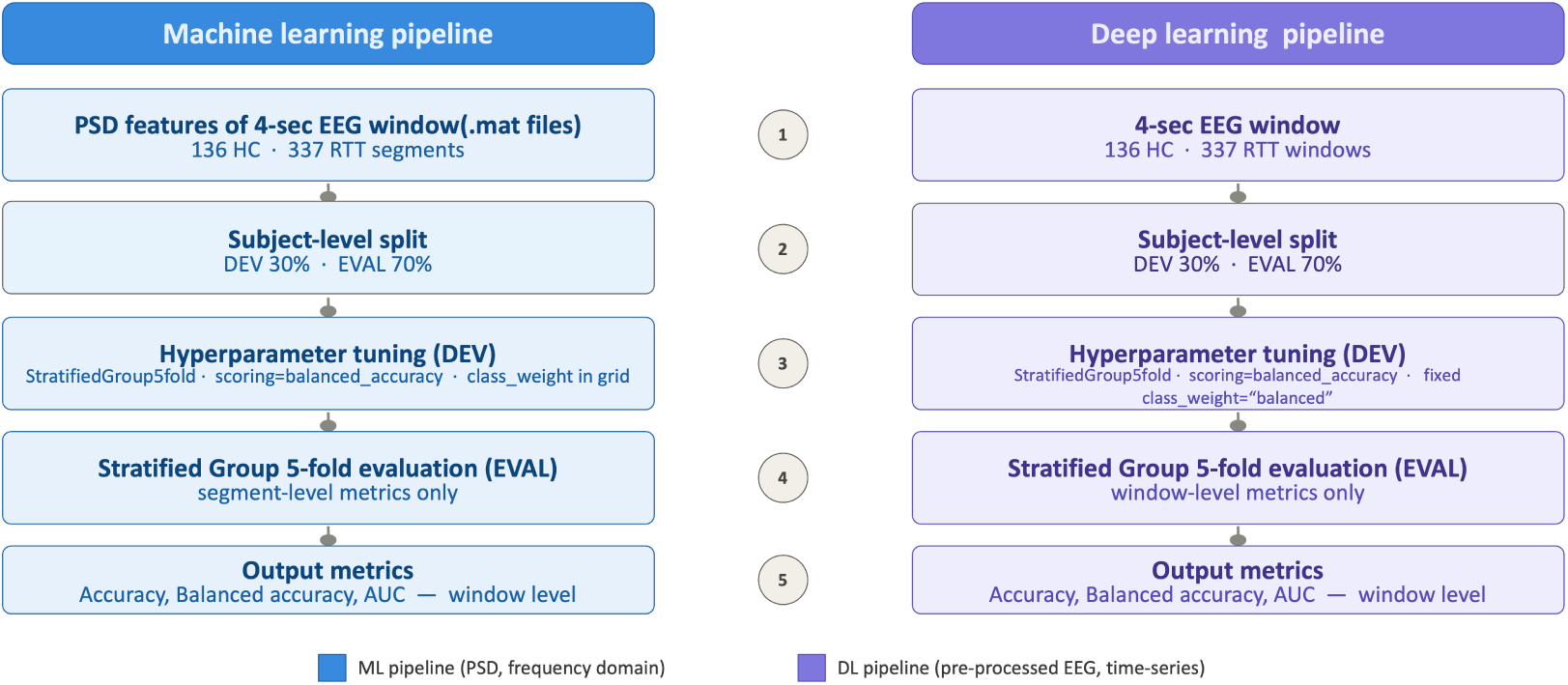
Overview of the machine learning and deep learning EEG classification pipelines used in this study.

To enhance model performance and avoid subject-level data leakage, the dataset was split into a development (DEV) set (30%; HC: RTT = 33:80) for hyperparameter tuning and an evaluation (EVAL) set (70%; HC: RTT = 103:257) for the final assessment. Hyperparameter tuning used Stratified Group k-fold cross-validation (*k* = 5) on the DEV set, ensuring recordings from the same participant remained in one fold while maintaining the class distribution. The best hyperparameters were then tested on the independent EVAL set using the same cross-validation approach.

Since the dataset exhibited class imbalance, balanced accuracy was used as the scoring metric for hyperparameter tuning. Cost-sensitive learning was also incorporated during model training. For classical ML models, class-weight hyperparameters were tuned where available, while XGBoost addressed imbalance using the scale_pos_weight parameter. Gradient Boosting does not support class weighting and was trained without explicit balancing. For DL models, class weights derived from the training data were incorporated into the loss function during optimisation.

For the DL pipeline, each EEG epoch was normalised using z-score standardisation prior to training. The models were optimised with the Adam optimiser, incorporating early stopping and dropout to enhance generalisation and prevent overfitting.

Hyperparameters tailored to each model were chosen solely based on the DEV set to maintain an unbiased assessment.

The final performance was evaluated using metrics such as accuracy, precision, sensitivity, specificity, balanced accuracy, F1-score, and the area under the receiver operating characteristic curve (AUC). These performance metrics were calculated for all EEG epochs in each test fold and then averaged over the five evaluation folds.

All analyses were implemented in Python (v3.10.20) within Jupyter Notebook. Data processing and machine learning experiments were conducted using NumPy (v1.26.4), pandas (v2.3.0), and scikit-learn (v1.7.2). Deep learning models were implemented using TensorFlow (v2.20.0), Keras (v3.11.3), and scikeras (v0.13.0). Matplotlib (v3.10.8) was used for visualising the results. To improve reproducibility, a fixed random seed (42) was used across all stochastic processes, including dataset splitting, cross-validation, and hyperparameter optimisation. Random seeds for TensorFlow and NumPy were also set for DL experiments, and all analyses were performed using singlethreaded evaluation (n_jobs=1). However, minor differences may still arise due to the inherent non-determinism of some DL operations.

### 2.8 Feature Importance Analysis

To enhance model interpretability and identify neurophysiologically meaningful EEG biomarkers, we conducted feature-importance analyses on the best-performing classical ML and DL models. We used separate explainability methods, each suited to their respective feature representations.

For the classical ML pipeline, SHapley Additive exPlanations (SHAP) analysis [51] was used on the top-performing model, Random Forest, to assess how each PSD feature impacts the model’s predictions. SHAP values were computed for all PSD-based features, each aligned with a specific channel-frequency bin. The absolute SHAP values were averaged across all test samples from the cross-validation folds to produce a global importance score for each feature. These scores were then mapped onto the original spectral-spatial data representation to derive channel-level and frequencyband importance, channel × frequency heatmaps, and scalp topographic maps, enabling interpretation of the key EEG channels and frequency regions that contributed to the classification.

For the DL pipeline, occlusion sensitivity analysis [52, 53] was applied to the best-performing model, EEGNet, which extracts representations directly from pre-processed EEG signals. Three complementary analyses were performed: (i) channel occlusion, in which individual EEG channels were masked; (ii) frequency-band occlusion, in which canonical EEG bands (delta, theta, alpha, beta, and gamma) were sequentially attenuated across all channels using a fourth-order Butterworth band-stop filter; and (iii) channel x band occlusion, in which specific frequency bands were attenuated within each EEG channel to produce a spatial-spectral importance matrix comparable to the SHAP-derived channel × frequency maps. Feature importance was quantified as the decrease in balanced accuracy relative to the original, unaltered model. Importance scores were visualised using bar plots, heatmaps, and scalp topographic maps.

Overall, these complementary explainability analyses enabled a direct comparison of the spatial and spectral EEG biomarkers identified by both classical ML and DL models.

## 3. Results

### 3.1 Machine Learning Classification Performance

Table 2 presents the performance of classical ML models using PSD features extracted from resting-state eyes-closed EEG. The performance metrics are reported as the mean *±* standard deviation across the five cross-validation folds.

**Table 2:** Classification performance of machine learning models on power spectral density features. Values represent the mean *±* standard deviation across five outer evaluation folds. The model with the best balanced accuracy (Bal. Acc.) is highlighted.

| Model | Performance Metrics (mean ± std) | | | | | | |
| --- | --- | --- | --- | --- | --- | --- | --- |
|  | Accuracy | Precision | Sensitivity | Specificity | F1 | Bal. Acc. | AUC |
| SVM (linear) | 0.782 ± 0.071 | 0.798 ± 0.084 | 0.839 ± 0.169 | 0.606 ± 0.331 | 0.706 ± 0.084 | 0.723 ± 0.099 | 0.855 ± 0.073 |
| SVM (RBF) | 0.819 ± 0.085 | 0.818 ± 0.098 | 0.853 ± 0.159 | 0.687 ± 0.247 | 0.768 ± 0.099 | 0.770 ± 0.096 | 0.858 ± 0.139 |
| Logistic Regression | 0.736 ± 0.096 | 0.754 ± 0.039 | 0.713 ± 0.215 | 0.735 ± 0.312 | 0.678 ± 0.095 | 0.724 ± 0.088 | 0.780 ± 0.094 |
| k-NN | 0.726 ± 0.182 | 0.725 ± 0.190 | 0.793 ± 0.223 | 0.529 ± 0.266 | 0.665 ± 0.180 | 0.661 ± 0.177 | 0.761 ± 0.214 |
| Decision Tree | 0.846 ± 0.051 | 0.833 ± 0.046 | 0.878 ± 0.101 | **0.735 ± 0.203** | 0.805 ± 0.061 | 0.807 ± 0.077 | 0.845 ± 0.091 |
| Random Forest | **0.885 ± 0.072** | 0.882 ± 0.090 | 0.939 ± 0.062 | 0.721 ± 0.273 | **0.837 ± 0.117** | **0.830 ± 0.127** | 0.933 ± 0.072 |
| Gradient Boosting | 0.866 ± 0.065 | 0.873 ± 0.077 | 0.951 ± 0.047 | 0.625 ± 0.254 | 0.803 ± 0.107 | 0.788 ± 0.119 | 0.930 ± 0.062 |
| XGBoost | 0.871 ± 0.061 | **0.898 ± 0.053** | **0.955 ± 0.059** | 0.637 ± 0.278 | 0.808 ± 0.109 | 0.796 ± 0.122 | **0.960 ± 0.033** |

Overall, tree-based ensemble methods consistently outperformed linear and distance-based classifiers. Random Forest achieved the highest balanced accuracy (0.830 ± 0.127) and F1-score (0.837 ± 0.117), along with a high AUC (0.933 ± 0.072), demonstrating the best overall balance between sensitivity and specificity, and an excellent ability to distinguish RTT from HC recordings. Meanwhile, XGBoost recorded the highest accuracy (0.871 ± 0.061), AUC (0.960 ± 0.033), precision (0.898 ± 0.053), and sensitivity (0.955 ± 0.059), though its lower specificity (0.637 ± 0.278) resulted in a slightly reduced balanced accuracy (0.796 ± 0.122). These results show that XGBoost was especially effective at detecting RTT recordings, correctly identifying the majority of positive cases.

Gradient Boosting and Decision Tree also showed strong performance. Gradient Boosting had high sensitivity (0.951 ± 0.047) but lower specificity (0.625 ± 0.254), while Decision Tree achieved the highest specificity (0.735 ± 0.203) among all classifiers, leading to a balanced accuracy similar to the top ensemble models.

Among the classifiers tested, the RBF SVM consistently outperformed both the linear SVM and Logistic Regression, indicating that nonlinear decision boundaries are advantageous for RTT classification. The kNN classifier showed the weakest results across all evaluation metrics.

Across all models, sensitivity was higher than specificity, suggesting the models were generally more effective at identifying RTT recordings than HCs. Overall, these results show that ensemble tree-based methods are especially effective at capturing the complex spectral features of EEG. Among them, Random Forest offered the best trade-off between sensitivity and specificity, while XGBoost demonstrated the strongest overall discriminative power, as indicated by the AUC.

### 3.2 Deep Learning Classification Performance

Table 3 presents the performance of DL architectures evaluated on preprocessed 4-second EEG segments. Results are presented as the mean ± standard deviation across five cross-validation folds.

**Table 3:** Classification performance of deep learning architectures on preprocessed 4-second EEG windows. Values represent mean *±* standard deviation across five outer evaluation folds. The best balanced accuracy (Bal. Acc.) is highlighted.

| Model | Performance Metrics (mean ± std) | | | | | | |
| --- | --- | --- | --- | --- | --- | --- | --- |
|  | Accuracy | Precision | Sensitivity | Specificity | F1 | Bal. Acc. | AUC |
| ShallowConvNet | 0.896 ± 0.049 | 0.887 ± 0.055 | 0.903 ± 0.072 | 0.857 ± 0.186 | 0.870 ± 0.062 | 0.880 ± 0.081 | 0.900 ± 0.127 |
| DeepConvNet | 0.813 ± 0.113 | 0.791 ± 0.128 | 0.813 ± 0.161 | 0.783 ± 0.270 | 0.773 ± 0.141 | 0.798 ± 0.136 | 0.868 ± 0.142 |
| EEGNet | **0.922 ± 0.041** | **0.921 ± 0.045** | **0.930 ± 0.066** | **0.870 ± 0.188** | **0.898 ± 0.063** | **0.900 ± 0.081** | **0.924 ± 0.100** |
| EEGConformer | 0.773 ± 0.109 | 0.706 ± 0.221 | 0.976 ± 0.015 | 0.279 ± 0.358 | 0.590 ± 0.204 | 0.627 ± 0.174 | 0.744 ± 0.184 |

Overall, EEGNet achieved the highest performance, with the highest accuracy (0.922 ± 0.041), precision (0.921 ± 0.045), F1-score (0.898 ± 0.063), and balanced accuracy (0.900 ± 0.081), along with a high AUC (0.924 ± 0.100). It also effectively balanced sensitivity (0.930 ± 0.066) and specificity (0.870 ± 0.188). ShallowConvNet performed similarly, with a balanced accuracy of 0.880 ± 0.081, an AUC of 0.900 ± 0.127, and specificity of 0.857 ± 0.186.

DeepConvNet showed moderate performance across all metrics, while EEGConformer had the lowest overall results. Although EEGConformer achieved the highest sensitivity (0.976 ± 0.015), its significantly lower specificity (0.279 ± 0.358) led to the lowest balanced accuracy (0.627 ± 0.174), suggesting it tends to overpredict the RTT class.

Across all architectures, sensitivity generally outperformed specificity, indicating that RTT recordings were easier to detect than HCs. However, EEGNet and ShallowConvNet offered a notably more balanced performance between these metrics compared to other models. Overall, the results show that EEG-specific DL models, especially EEGNet, provide reliable end-to-end classification of RTT from preprocessed EEG data and offer a benchmark for comparison with traditional ML approaches.

### 3.3 Identification of EEG Biomarkers Using Explainable AI

#### 3.3.1 SHAP Analysis of the Random Forest Model

Figure 4a illustrates the spatial and spectral distribution of SHAP importance across EEG frequency bands. Delta and theta activities showed the highest importance in the left centro-frontal region, particularly around C3, whereas alpha and beta contributions were more widespread across frontocentral and posterior areas. In contrast, gamma activity showed relatively weak and localised importance over posterior electrodes. The overall topographic map confirmed that the most distinctive EEG features were concentrated over the left centro-frontal cortex, with less significant contributions from the bilateral frontal and central regions.

**Figure 4:**
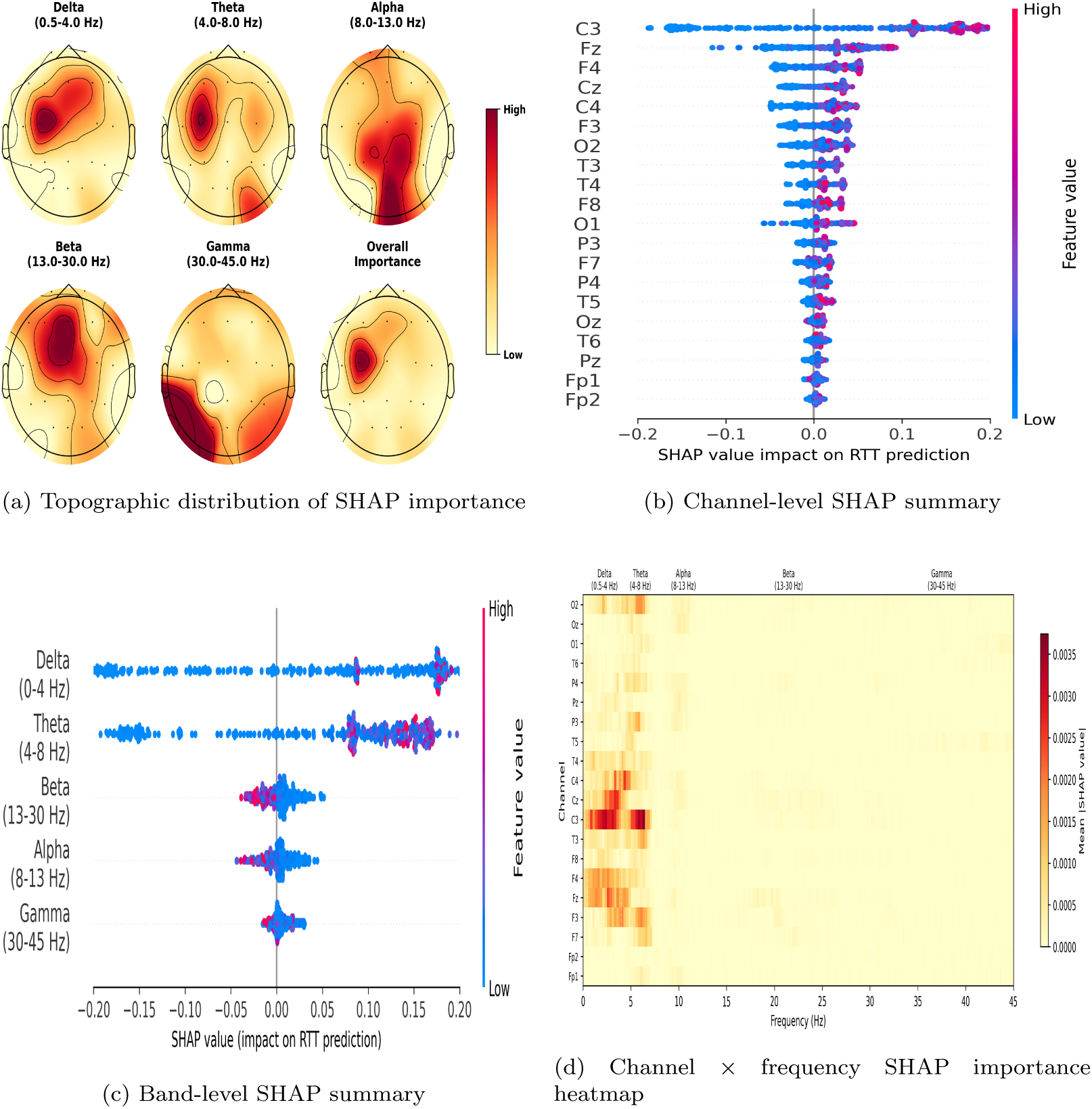
SHAP-based interpretation of the Random Forest classifier. (A) Spatial distribution of channel importance across EEG frequency bands together with overall channel importance. (B) Channel-level SHAP summary showing the influence of each EEG channel on model predictions. (C) SHAP summary aggregated by frequency band. (D) Channel × frequency SHAP importance heatmap illustrating the joint spatial and spectral contribution of EEG features. Warmer colours indicate greater feature importance.

Consistent with these results, the channel-level SHAP summary (Figure 4b) identified C3 as the most influential electrode for RTT classification, followed by Fz, F4, Cz, C4, F3, O2, and T3. Frontal-polar (Fp1 and Fp2) and posterior midline electrodes contributed minimally. The channel x frequency SHAP heatmap (Figure 4d) further revealed that the model predominantly relied on low-frequency spectral components, with the highest SHAP values observed in the delta (0.5–4 Hz) and theta (4–8 Hz) bands over C3 and nearby fronto-central electrodes. Importance sharply declined above approximately 10 Hz, suggesting limited influence of higher-frequency oscillations.

Band-level SHAP analysis (Figure 4c) confirmed that delta and theta were the most influential contributors to the model’s predictions, while alpha, beta, and gamma had comparatively smaller impacts. The SHAP summary plot revealed that higher delta and theta power tended to steer predictions towards the RTT class (red points), whereas lower values favored the HC class (blue points). Overall, these results show that the Random Forest classifier mainly based its decisions on low-frequency activity (cortical slowing) originating from left centro-frontal brain regions for RTT detection.

#### 3.3.2 Occlusion Sensitivity Analysis of EEGNet

Channel occlusion analysis (see Figure 5b) identified C3 as the key electrode, with F3, Oz, F4, C4, and Fz also playing significant roles. This indicates that EEGNet mainly relied on centro-frontal and posterior brain regions for RTT classification. Conversely, blocking channels such as T4, Fp2, and Cz caused little or no drop in balanced accuracy, indicating that these electrodes provided limited discriminative information.

**Figure 5:**
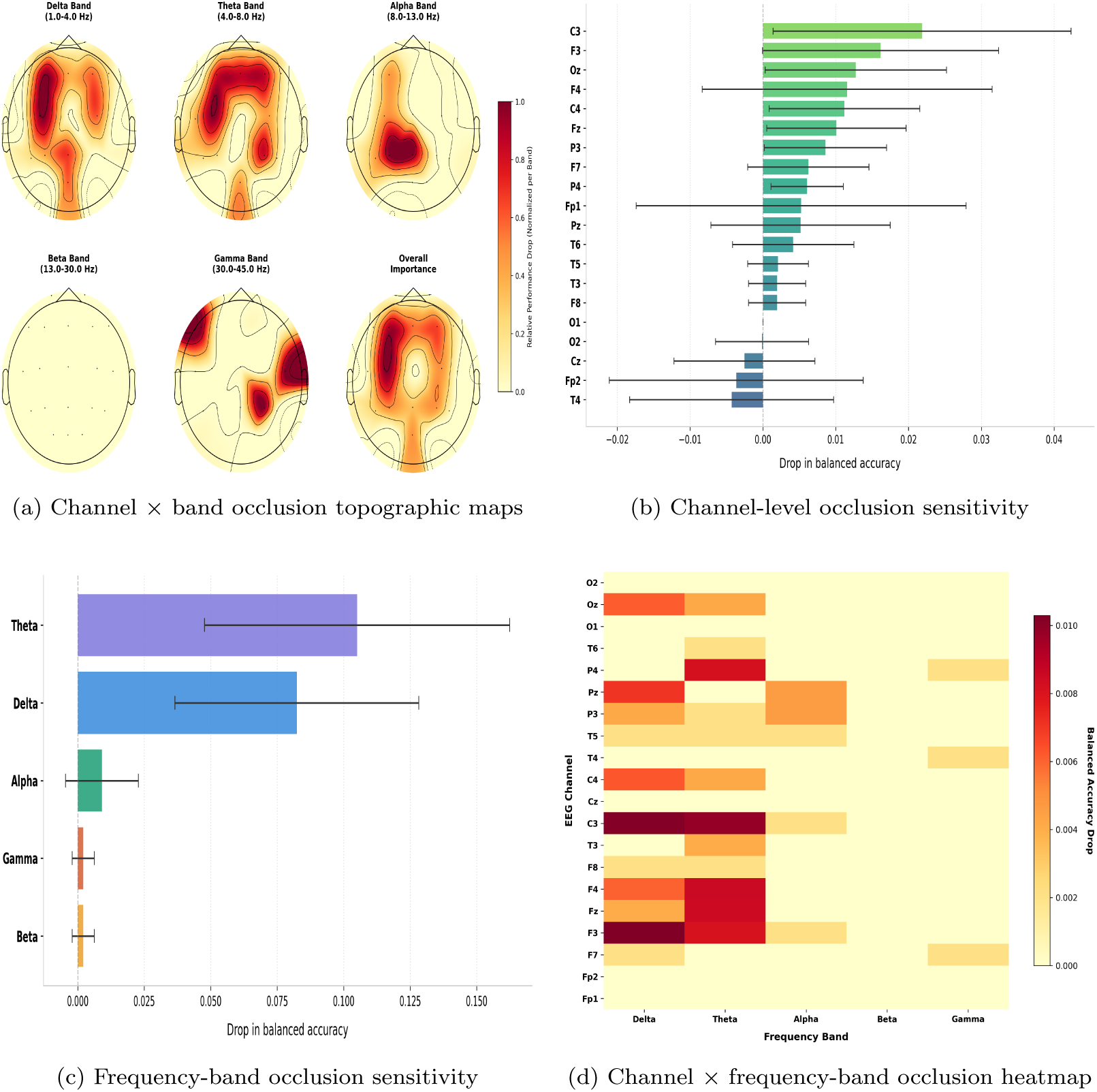
Occlusion sensitivity analysis of the EEGNet classifier. (A) Topographic distribution of channel importance for each canonical EEG frequency band; the overall map represents the mean importance across all bands. (B) Channel-level occlusion sensitivity showing the reduction in balanced accuracy following occlusion of each EEG channel. (C) Frequency-band occlusion sensitivity showing the reduction in balanced accuracy following attenuation of each canonical EEG frequency band. (D) Channel × frequency-band occlusion heatmap illustrating the joint spatial and spectral importance learned by EEGNet. Larger reductions in balanced accuracy indicate greater feature importance.

Frequency-band occlusion (Figure 5c) showed that the theta band was the most informative, as its removal caused the largest drop in balanced accuracy (0.105 ± 0.058), with the delta band causing a slightly smaller decline (0.083 ± 0.046). In contrast, blocking the alpha, beta, and gamma bands led to only minor performance changes (<0.01), suggesting that higher-frequency oscillations contribute less.

The channel × band occlusion heatmap (Figure 5d) showed that the most pronounced performance declines occurred mainly in the delta and theta bands across electrodes C3, F3, F4, C4, Pz, and Oz. By contrast, alpha-band effects were weaker and largely confined to centro-parietal electrodes. Occluding the beta and gamma bands produced little to no change across most channels. These findings were further supported by the band-specific topographic maps (Figure 5a), which indicated that delta and theta activity was most prominent over centro-frontal areas, particularly the left central cortex. In contrast, contributions from the alpha, beta, and gamma bands were weaker and more localised to central and peripheral electrodes. Overall, the occlusion analysis suggests that EEGNet primarily relies on low-frequency activity from centro-frontal cortical regions to distinguish RTT from healthy controls.

## 4. Ablation Study: Effect of Epoch Length on Classification Performance

The impact of EEG epoch length on classification performance was assessed using 2-, 4-, and 6-second windows for both traditional machine learning and deep learning models. The performance results are shown in Figures 6 and 7 depicting variations in balanced accuracy and AUC compared to the 4-second baseline.

**Figure 6:**
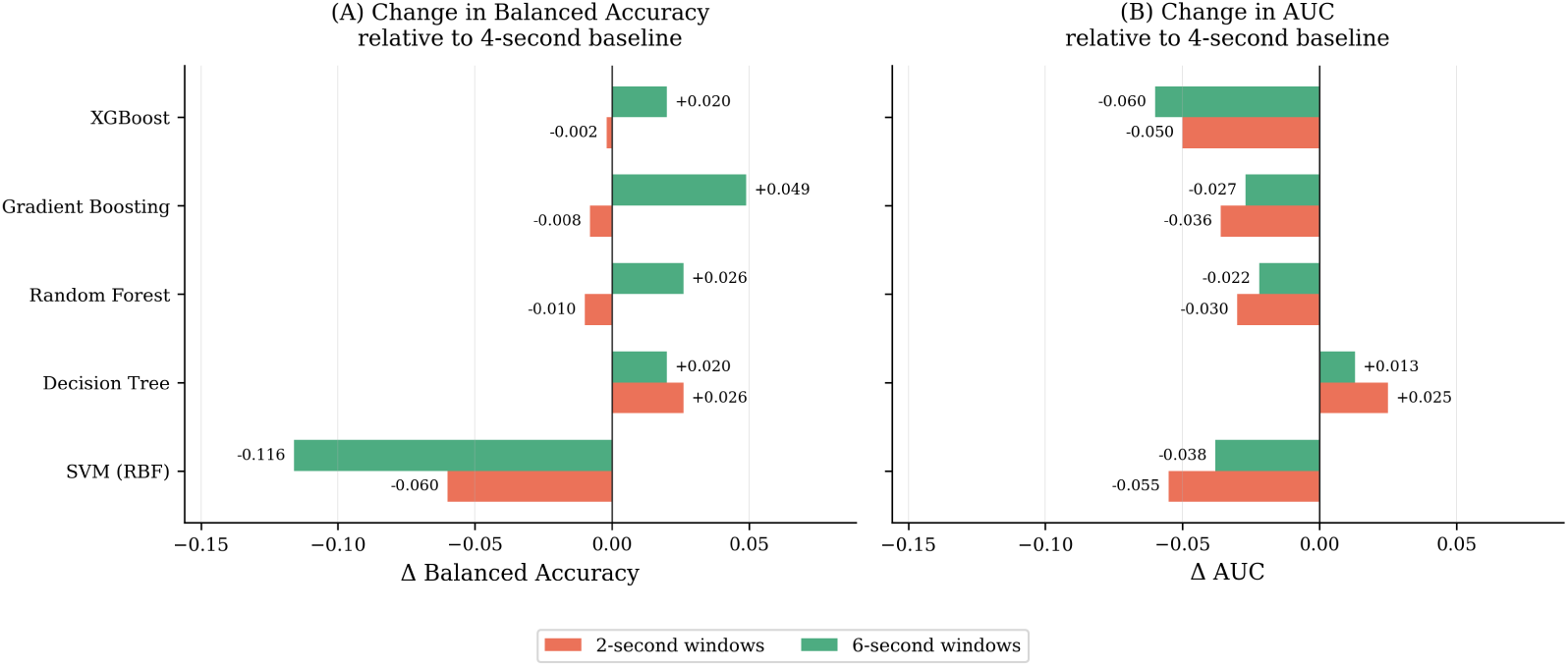
Ablation study: Effect of Epoch Length on Classification Performance of top five best performing machine learning algorithms

**Figure 7:**
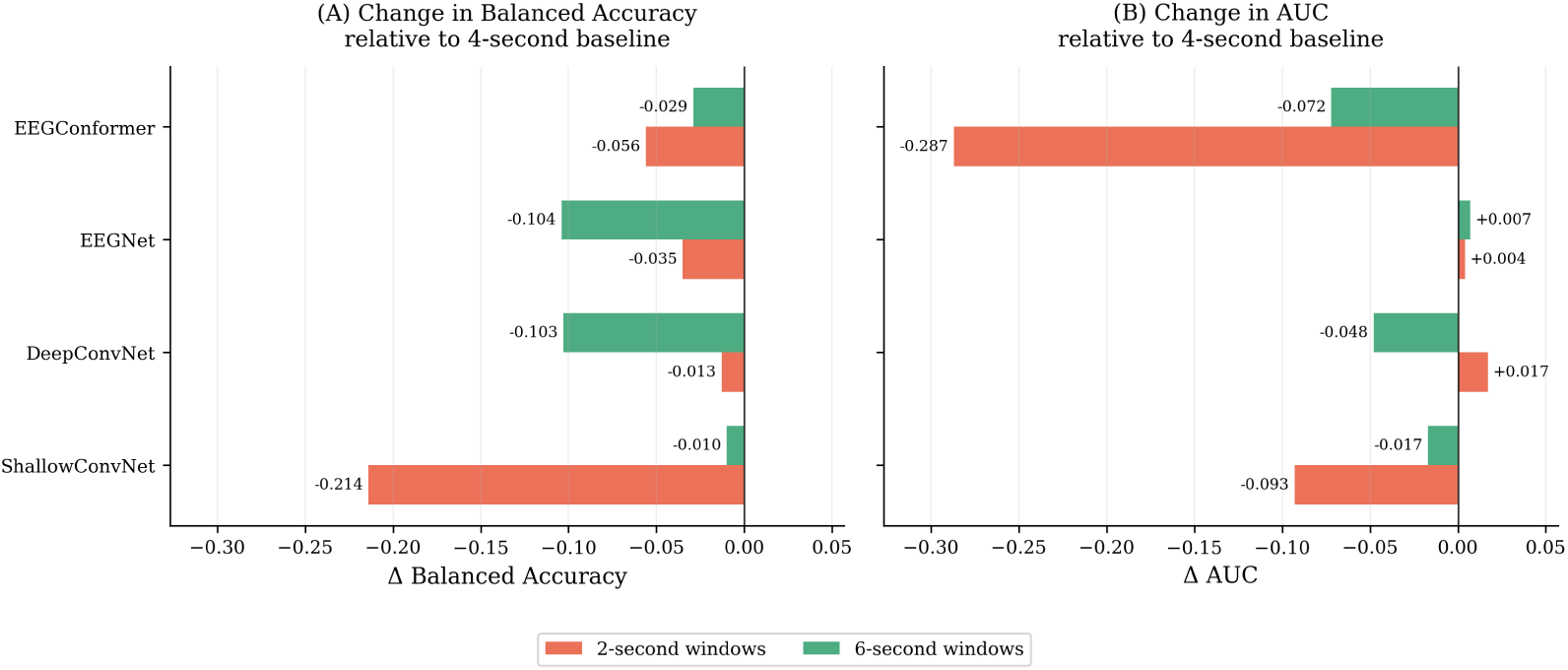
Ablation study: Effect of Epoch Length on Classification Performance of deep learning algorithms

Among the classical ML models, Random Forest achieved the highest balanced accuracy across both the 4and 6-second configurations. Decision Tree performed best with 2-second windows. Compared with the 4-second baseline, 6-second windows slightly increased Random Forest’s balanced accuracy from 0.830 to 0.856 and caused only a small drop in AUC from 0.933 to 0.911. On the other hand, 2-second windows led to a minor decrease in balanced accuracy (0.833) and AUC (0.870). Among all evaluated models, Gradient Boosting showed the greatest improvement with 6-second windows, while SVM (RBF) experienced the most significant performance decline at both shorter and longer durations.

For the DL models, EEGNet consistently achieved the highest performance with 4-second windows, yielding a balanced accuracy of 0.900 and an AUC of 0.924. Both shorter and longer epochs reduced classification accuracy. Using 2-second windows decreased the balanced accuracy to 0.865, while 6-second windows further reduced it to 0.811. Similar patterns were observed with DeepConvNet and EEGConformer, although ShallowConvNet showed the largest decline in 2-second windows, with a 21.4% reduction in balanced accuracy relative to the baseline. Overall, the 4-second epoch offered the optimal classification performance among the evaluated deep learning configurations.

## 5. Discussion

### 5.1 Main Findings

This study shows that both classical ML and DL can accurately differentiate RTT from HC using resting-state, eyes-closed EEG recordings and identify key physiological biomarkers. Among the evaluated methods, Random Forest achieved the highest accuracy and balanced accuracy among classical ML models, while EEGNet showed the best overall performance among DL architectures. These findings suggest that both handcrafted frequency features and end-to-end learning from preprocessed EEG data are effective for RTT classification. Additionally, the ablation study indicated that 4-second EEG epochs provide the best balance between classification accuracy and temporal context for both modelling approaches.

A key insight from this study is the strong consensus between the explainability analyses of the two modelling approaches. Despite different feature representations, SHAP analysis of the Random Forest classifier and occlusion sensitivity analysis of EEGNet both consistently highlighted low-frequency oscillations, particularly in the delta and theta bands, and centro-frontal cortical regions as the main contributors to RTT classification. The agreement across these independent explainability methods indicates that these biomarkers genuinely reflect RTT-related neurophysiological abnormalities rather than artefacts specific to one model.

Compared with previous RTT EEG research, this study advances the field by being, to our knowledge, the first to systematically compare PSD-based classical ML with end-to-end DL within a unified evaluation framework. It not only confirms the viability of modern EEG-tailored DL models for RTT classification but also shows that independent explainability methods consistently identify similar spatial and spectral biomarkers. This enhances trust in the biological significance of these EEG features and offers a clearer, more transparent AI-based RTT diagnosis framework beyond prediction accuracy.

Overall, these results show that high diagnostic performance can be achieved while maintaining interpretability. By combining rigorous evaluation with additional explainability methods, the proposed framework provides a foundation for developing reliable, clinically relevant EEG biomarkers for Rett syndrome and for future research investigating disease severity, patient stratification, and treatment monitoring.

### 5.2 Comparative Analysis of Classical Machine Learning and Deep Learning Models

This paper compares classical ML and DL methods for automated RTT classification using resting-state, eyes-closed EEG data. To this end, we evaluated PSD-based classical ML models against DL architectures trained directly on preprocessed EEG windows.

Among classical ML models, ensemble tree-based methods consistently achieved the strongest performance, suggesting that the relationship between EEG spectral features and RTT is nonlinear and involves complex interactions among channels and frequency components. The superior performance of Random Forest and XGBoost relative to Logistic Regression, linear SVM, and k-nearest neighbours, together with the improvement of the RBF SVM over the linear SVM, supports this interpretation. Additionally, the highest AUC achieved in this study (0.960) is comparable to those reported in previous RTT EEG studies using resting-state spectral features (AUC *∼* 0.92) [16] and auditory event-related potentials (AUC *∼* 0.94) [19], further supporting the strong discriminative value of resting-state eyes-closed PSD features.

Among deep learning models, EEGNet achieved the highest overall accuracy, while ShallowConvNet, despite its simpler design, achieved comparable performance. In contrast, deeper models such as DeepConvNet and the transformer-based EEGConformer did not improve classification accuracy, suggesting that increased architectural complexity does not necessarily yield better performance on relatively small RTT datasets. The strong performance of ShallowConvNet, which was intentionally designed to mimic traditional EEG band-power extraction, highlights that spectral information remains a highly valuable feature for RTT classification.

A direct comparison of the best-performing models (see Table 4) shows that EEGNet achieved the highest balanced accuracy, accuracy, specificity, precision, and F1-score, whereas the Random Forest model recorded the highest sensitivity and AUC. These findings indicate that an end-to-end deep learning model provides a modest improvement in overall classification performance, although handcrafted PSD features remain highly competitive for differentiating RTT from healthy controls.

**Table 4:** Performance comparison between the best-performing machine learning and deep learning models based on the balanced accuracy. Values represent the mean *±* standard deviation across five outer evaluation folds.

| Pipeline | Best Model | Accuracy | Precision | Sensitivity | Specificity | F1 | Bal. Acc. | AUC |
| --- | --- | --- | --- | --- | --- | --- | --- | --- |
| ML (PSD features) | Random Forest | 0.885 ± 0.072 | 0.882 ± 0.090 | **0.939 ± 0.062** | 0.721 ± 0.273 | 0.837 ± 0.117 | 0.830 ± 0.127 | **0.933 ± 0.072** |
| DL (EEG windows) | EEGNet | **0.922 ± 0.041** | **0.921 ± 0.045** | 0.930 ± 0.066 | **0.870 ± 0.188** | **0.898 ± 0.063** | **0.900 ± 0.081** | 0.924 ± 0.100 |

The enhanced performance of EEGNet stems from its ability to learn hierarchical spatial and temporal features directly from preprocessed EEG signals [49], capturing temporal dynamics and spatial correlations across EEG channels using convolutional filters tailored to electrophysiological data. This end-to-end learning approach enables the neural network to identify subtle discriminative patterns that spectral power alone might miss, leading to improvements in balanced accuracy, specificity, and F1-score.

In contrast, the Random Forest classifier achieved competitive performance using only PSD features, indicating that spectral information captures much of the discriminative signal associated with RTT. Moreover, the explicit PSD features enable direct physiological interpretation, providing insights into the spectral factors that influence classification.

The fold-wise evaluation (Table 5) further highlights that both models consistently exhibited high sensitivity across all evaluation folds, demonstrating reliable RTT detection. While their performances were comparable in most folds, EEGNet achieved higher balanced accuracy on the most difficult partition (fold 2) and exhibited less variability across folds than Random Forest, indicating more stable generalisation.

**Table 5:** Per-fold classification performance for the best-performing ML model (Random Forest) and DL model (EEGNet) across the five outer evaluation folds. Balanced accuracy, AUC, and Sensitivity are reported per fold to illustrate foldto-fold variability.

| Fold | ML - Random Forest |  |  | DL - EEGNet |  |  |
| --- | --- | --- | --- | --- | --- | --- |
|  | Bal. Acc. | AUC | Sensitivity | Bal. Acc. | AUC | Sensitivity |
| 1 | 0.750 | 0.812 | 1.000 | 0.750 | 0.728 | 1.000 |
| 2 | 0.621 | 0.889 | 0.941 | 0.971 | 0.975 | 0.941 |
| 3 | 0.971 | 1.000 | 1.000 | 0.971 | 0.998 | 1.000 |
| 4 | 0.891 | 0.972 | 0.918 | 0.893 | 0.935 | 0.878 |
| 5 | 0.917 | 0.993 | 0.833 | 0.917 | 0.983 | 0.833 |
| **Mean** | 0.830 | 0.933 | 0.939 | 0.900 | 0.924 | 0.930 |
| **Std** | 0.127 | 0.072 | 0.062 | 0.081 | 0.100 | 0.066 |

Overall, these results show that neither is universally superior. Classical ML offers better interpretability, lower computational demands, and strong diagnostic accuracy, while DL improves overall classification by utilising EEG waveform data that engineered spectral features do not explicitly capture. These results support DL for optimal diagnostic accuracy while affirming that well-designed spectral features remain a valuable and interpretable option for RTT classification, especially when data is limited.

### 5.3 Neurophysiological interpretation of the identified EEG biomarkers

In addition to evaluating classification performance, this study aims to identify neurophysiologically relevant EEG biomarkers that reliably distinguish RTT from HC. To this end, additional XAI analyses were conducted using SHAP with the Random Forest classifier and occlusion sensitivity analysis with EEGNet, enabling a comparison of biomarkers identified by classical ML and DL approaches.

The findings show that the EEG biomarkers most closely linked to Rett syndrome are low-frequency oscillations, particularly delta and theta waves, predominantly localised to the centro-frontal cortical regions. Although the two analyses used different feature representations, they identified similar spatial and spectral patterns. This consistent overlap indicates that the biomarkers reflect true neurophysiological abnormalities associated with RTT rather than model-specific artefacts.

Both explainability analyses consistently identified delta and theta activity as the main factors influencing classification. This aligns with prior research indicating generalised cortical slowing in RTT, characterised by increased delta and fronto-central theta activity along with decreased posterior alpha rhythms [14, 16]. These abnormalities are associated with impaired cortical development, disrupted excitation–inhibition balance, and altered thalamocortical communication resulting from MECP2 dysfunction [54, 55, 56, 57]. While reduced alpha power is a recognised RTT feature, both SHAP and occlusion analyses suggest that pathological increases in slow-wave activity provide more robust discriminative information than changes in the alpha, beta, or gamma bands.

The spatial distribution of feature importance further supports this interpretation. Both SHAP topographic maps and occlusion sensitivity maps consistently highlight the centro-frontal cortex, particularly around electrodes C3, F3, F4, Fz, and neighbouring electrodes, with smaller contributions from occipital regions. These regions are involved in sensorimotor integration, motor planning, and higher cognitive functions, which are known to be affected in RTT [58]. Similarly, channel x frequency analyses show that the most informative features are predominantly below 8 Hz over these centro-frontal electrodes, with importance dropping significantly in the alpha, beta, and gamma bands. Although SHAP and occlusion measure feature importance differently, their strong agreement in spatial and spectral patterns suggests that both Random Forest and EEGNet rely on similar neurophysiological signatures of RTT.

An additional advantage of SHAP analysis is its ability to offer directional insights into model predictions. Elevated delta and theta power primarily drive predictions toward the RTT class, while reduced slow-frequency power is associated with healthy controls.

In summary, the common findings between SHAP and occlusion sensitivity analyses suggest that both traditional machine learning and deep learning models learn biologically meaningful EEG features consistent with the known pathophysiology of RTT. These findings enhance the transparency of the diagnostic framework and suggest that low-frequency centro-frontal EEG abnormalities could serve as objective biomarkers to automate RTT diagnosis.

### 5.4. Influence of EEG epoch length on classification performance

The ablation study shows that EEG epoch length is a key factor in automated RTT classification, affecting both the quality of EEG data and how well ML models can generalise. While the ideal epoch length varies with the modelling method, current results suggest that moderate-length epochs strike the best balance between capturing signals effectively and maintaining sufficient data.

For classical ML models, the stability of spectral features is crucial to performance. Short epochs provide limited frequency resolution, particularly for low-frequency oscillations such as delta and theta activity, which are key biomarkers of RTT identified in the explainability analyses. In contrast, longer epochs improve spectral estimation but reduce the number of independent training samples, thereby limiting the feature space the model can learn from. These results indicate that an intermediate epoch duration yielded the most consistent results.

The deep learning models showed a similar overall trend, despite learning directly from EEG waveforms rather than handcrafted features. Deep neural networks require sufficient temporal context to develop meaningful spatial and temporal representations, but very long epochs reduce the available training data for optimisation. This trade-off is particularly critical in RTT, where the disorder’s rarity naturally limits dataset size. The consistent performance of EEGNet across various epoch lengths indicates that compact EEG-specific architectures are more robust to changes in temporal resolution than deeper convolutional or transformer-based models.

Overall, these findings suggest that epoch length should be considered a configurable model parameter rather than a fixed preprocessing setting. While longer windows enhance frequency resolution and shorter ones increase the number of training samples, neither extreme consistently yields better classification results. The findings indicate that a 4-second window strikes a good balance between capturing meaningful spectral information, providing sufficient temporal context for deep learning models, and maximising the number of independent training examples. Achieving this balance is especially crucial for small clinical EEG datasets and can serve as a practical guideline for future RTT research and other rare neurodevelopmental disorders.

### 5.5. Limitations and Future Work

This study shows that both PSD-based machine learning and end-to-end deep learning can effectively classify Rett syndrome and identify EEG biomarkers that are physiologically relevant. Although the classification results are promising and the interpretability findings are consistent, several limitations warrant acknowledgment.

First, the study was limited by its relatively small sample size, reflecting the rarity of Rett syndrome, and by an imbalanced class distribution, with fewer HC than the RTT cohort. Furthermore, the RTT and control groups were not fully matched for age and sex, a common challenge in rare-disorder studies, where recruiting matched controls is often difficult. Despite efforts to reduce bias through balanced accuracy as the primary evaluation metric, class-weighted model training, and subject-wise nested cross-validation, these cohort differences may have affected the results and limited the generalisability of the findings.

Second, the analysis focused solely on resting-state EEG recordings with eyes closed. Although this ensures a stable physiological state and reduces task-related confounding factors, it does not detect abnormalities that may arise during eyes-open recordings, cognitive tasks, or event-related paradigms. Third, different explainability methods were required for the two modelling approaches. SHAP was used with the PSD-based ML model, whereas occlusion sensitivity analysis was employed for the DL model, given its endto-end feature representation. Although both approaches identified highly consistent biomarkers, other deep learning explanation techniques, such as Layer-wise Relevance Propagation [59], EEG-specific Grad-CAM methods [60], and concept-based explanations [61], could offer additional insights into network functioning.

Finally, the framework was tested solely for binary classification between RTT and HC. As a result, the specificity of the biomarkers for distinguishing RTT from other neurodevelopmental disorders remains uncertain.

Future research should validate this framework using larger, ageand sexmatched, multi-centre cohorts and extend the analysis to include additional recording paradigms and multimodal datasets. Testing the models in multiclass settings involving disorders such as Angelman syndrome, CDKL5 deficiency disorder, and FOXG1 syndrome will help verify the specificity of the identified biomarkers. Additionally, longitudinal studies with larger cohorts, incorporating genotype data, clinical severity scores, and treatment outcomes, could help establish whether the identified EEG biomarkers are linked to disease progression, symptom severity, or response to therapy. These studies would support the development of interpretable AI systems that not only enable automatic diagnosis but also assist with patient stratification, prognosis, and monitoring of treatment effectiveness in Rett syndrome.

## 6. Conclusion

In this work, we demonstrate that artificial intelligence can achieve both diagnostic accuracy and physiological interpretability in EEG-based Rett syndrome classification. A key finding is that classical machine learning models (such as Random Forest, XGBoost, and Gradient Boosting) and deep learning models (such as ShallowConvNet and EEGNet), despite employing fundamentally different learning strategies, are not competing but complementary. Classical machine learning models offer transparent interpretation via explicit spectral feature representations, whereas deep learning architectures learn richer hierarchical representations directly from preprocessed EEG signals, thereby improving classification performance. Beyond diagnostic performance, this work emphasises the importance of integrating explainability into biomedical artificial intelligence. Both approaches, namely SHAP for machine learning models and occlusion sensitivity analysis for deep learning models, identified abnormal low-frequency activity in centro-frontal brain regions (particularly, C3, F3, F4, Fz) as key biomarkers for differentiating RTT from healthy controls, aligning with known RTT pathology. Such transparent, explainable models are more likely to gain clinical acceptance and to be integrated into standard diagnostic workflows than opaque black-box systems, providing clinicians with an understandable rationale for automated diagnoses.

This work establishes a benchmark for future EEG-based artificial intelligence research in Rett syndrome. To the best of our knowledge, this is the first study to systematically compare classical machine learning with deep learning for EEG-based RTT classification, and to include explainability analyses within a unified framework. Our findings show that high diagnostic accuracy can be combined with physiological interpretability, yielding reliable, transparent, and clinically applicable AI systems for RTT diagnosis and monitoring. Future research should explore whether the identified EEG biomarkers are associated with disease severity, genotype, or longitudinal clinical outcomes, enabling patient stratification, prognosis, and monitoring of therapeutic interventions in Rett syndrome.

## Data Availability

The code for training and evaluating the machine learning and deep learning classification models used in this study is publicly available on GitHub at https://github.com/meghag-git/eeg-classification-rtt.git.
Given the sensitive and identifying nature of the data, and in line with the consent agreed with participants, the data that support the findings of this study are not publicly available.

## Acknowledgment

The authors acknowledge Research Line 1 of the IRCCS Stella Maris Foundation (RC 2025), within the research activities supported by the Italian Ministry of Health, for its commitment to advancing research in the field of rare neurogenetic diseases. The authors also acknowledge Ms Laura Noble (MSc student, Trinity College Dublin) for her support and assistance with parts of the EEG data preprocessing pipeline.

## Funding

This publication has emanated from research supported under the European Union’s Horizon 2020 research and innovation programme under the Marie Skłodowska-Curie grant agreement No 101034252, with the financial support of Research Ireland [12/RC/2289_P2], at the Insight Research Ireland Centre for Data Analytics at University College Dublin and from research supported in part by a research grant from Research Ireland under Grant Number 21/RC/10294_P2 and co-funded under the European Regional Development Fund and by FutureNeuro industry partners.

## CRediT authorship contribution statement

**Megha Gupta:** Writing Original Draft, Writing Review & Editing, Conceptualization, Formal analysis, Methodology, Investigation, Visualization, Software, Funding acquisition. **Changhong Jin:** Writing Review & Editing, Validation. **Alejandro Lopez Valdes:** Writing Review & Editing, Validation. **Chiara Bosetti:** Writing Review & Editing, Data Curation. **Gemma Marinella:** Writing Review & Editing, Data Curation. **Emanuele Bartolini:** Writing Review & Editing, Data Curation, Resources. **Roberta Battini:** Data Curation, Resources, Funding acquisition. **Ruihai Dong:** Writing Review & Editing, Conceptualization, Project administration, Supervision, Funding acquisition. **Daniela Tropea:** Writing Review & Editing, Conceptualization, Data Curation, Project administration, Resources, Supervision, Funding acquisition.

## Code availability statement

The code for training and evaluating the machine learning and deep learning classification models used in this study is publicly available on GitHub at https://github.com/meghag-git/eeg-classification-rtt.git.

## Data availability

Given the sensitive and identifying nature of the data, and in line with the consent agreed with participants, the data that support the findings of this study are not publicly available.

## Declaration of competing interest

The authors declare that they have no known competing financial interests or personal relationships that could have appeared to influence the work reported in this paper.

## Generative AI disclosure statement

During the preparation of this work, the authors used ChatGPT (OpenAI) in order to assist with language editing, paraphrasing, and improving the clarity and readability of the manuscript. After using this tool, the authors reviewed and edited the content as needed and takes full responsibility for the content of the published article.

## Notes

### Competing Interest Statement

The authors have declared no competing interest.

### Author Declarations

The Pediatric Ethics Committee of the Tuscany Region (CEPR) of IRCCS Stella Maris Foundation in Pisa, Italy gave ethical approval for this work.

